# THE IMPACT OF THE MEDITERRANEAN DIET AND LIFESTYLE INTERVENTION ON LIPOPROTEIN SUB-CLASS PROFILES AMONG METABOLIC SYNDROME PATIENTS. FINDINGS OF A RANDOMIZED CONTROLLED TRIAL

**DOI:** 10.1101/2023.07.27.23293292

**Authors:** Beatriz Candás-Estébanez, Bárbara Fernández-Cidón, Emili Corbella, Cristian Tebé, Marta Fanlo-Maresma, Virginia Esteve-Luque, Jordi Salas-Salvadó, Montse Fitó, Antoni Riera-Mestre, Emilio Ros, Xavier Pintó

## Abstract

**Background:** Metabolic syndrome (MetS) is associated with alterations of lipoprotein structure and function that can be characterized with advanced lipoprotein testing (ADLT). The effect of Mediterranean diet (MedDiet) and of body weight loss on the lipoprotein subclass profile has been scarcely studied. Within the PREDIMED-Plus randomized controlled trial, a sub-study conducted in one of its recruiting centers aimed to evaluate the effects on lipoproteins properties assessed by ADLT of an intensive weight loss program based on an energy-reduced MedDiet (er-MedDiet) and physical activity (PA) promotion (intervention group) compared with energy-unrestricted MedDiet recommendations (control group) on lipoprotein subclasses.

**Methods:** 202 patients with MetS (n=107, Intervention; n=95, Control) were included in this study. Conventional lipid profile and ADLTs were performed at baseline, 6 months and one year. Linear mixed models were used to assess the effects of intervention on lipoprotein profiles.

**Results:** The er-MedDiet+PA led to a significant decrease of body mass index by 1.5 Kg/m^2^ at 6 months and 1.4 Kg/m^2^ at 12 months and a reduction of waist circumference by 2.9 cm at 6 months and 2.2 cm at 12 months; an increase of 0.1 mmol/L of HDL-C at 12 months, and decreased triglycerides (Tg) at 6 months; and decreased LDL-C and non-HDL-C at 12 months. ADLT showed a decrease of small dense-LDL-C (sdLDL-C), intermediate-density lipoprotein (IDL)-C, and HDL-Tg, and an increase of large LDL-particles (P). The er-MedDiet+PA model predicted significant reductions of Tg, sdLDL-C, VLDL-Tg and large VLDL-P in the intervention group.

**Conclusions:** In comparison with MedDiet (control group), er-MedDiet+PA (intervention group) decreased plasma triglyceride, and triglyceride content in HDL and VLDL particles, decreased sdLDL-C, and increased large LDL particles, indicating beneficial changes against cardiovascular disease.

## 1. Introduction

Cardiovascular disease (CVD) remains the leading cause of morbidity and mortality worldwide [1, 2]. The concentration of low-density lipoprotein cholesterol (LDL-C) is tightly linked to CVD mortality [3, 4] and is the main target of CVD prevention strategies [5]. However, ischemic events also do occur in populations with an LDL-C concentration below the cutoff value used to define increased cardiovascular risk (CVR) [6], mainly in patients with metabolic syndrome (MetS) [7]. MetS is a clinical condition with insulin resistance and central obesity leading to glucose intolerance, dyslipidemia and increased blood pressure as key components. Managing MetS requires modifying lifestyle habits, such as reducing weight through dieting and increasing physical activity [8]. Higher adherence to the Mediterranean diet (MedDiet) has a beneficial impact on lipid alterations and other components of the MetS [9,10] and is also associated with reduced mortality related to this disorder [11–13]. The PREDIMED trial has shown that the MedDiet has a protective effect against CVD [14,15]. This salutary effect can be attributed to the myriad beneficial nutrients and bio-actives contained in MedDiet foods, such as vegetable proteins, monounsaturated and polyunsaturated fatty acids, dietary fibre, vitamins, non-sodium minerals, and polyphenols [16], that are contained in extra-virgin olive oil, whole grains, nuts and a wide variety of fruits and vegetables that are characteristic foods of the MedDiet [17]. In this sense, it has been reported that the consumption of polyphenol-rich extra-virgin olive oil decreases the atherogenicity of LDL particles [18,19].

Each type of lipoprotein, including LDL, can be sorted into subclasses according to differing size, density, and composition. Because of their differing physical properties, the impact of each lipoprotein subclass on CVR is also different [20]. MetS patients have abnormal lipid profiles consisting of a “lipid triad” of: 1) increased triglycerides (Tg); 2) decreased high-density lipoprotein cholesterol (HDL-C); 3) small dense LDL (sdLDL) particles as the dominant subclass of LDL [21]. sdLDL particles are more atherogenic, in part because a higher number of sdLDL particles than large LDL (lLDL) particles is needed to carry the same amount of plasma cholesterol, and the higher the number of LDL particles, the higher the CVR [22]. sdLDL particles also promote atherosclerotic plaque rupture, which triggers ischemic events [23].

In the last years new methods that characterize lipoproteins more accurately have been developed, including advanced lipoprotein testing (ADLT) based on Nuclear Magnetic Resonance Spectroscopy (NMR)[24], and novel precipitation assays to directly quantify sdLDL-C. However, the effects of the MedDiet on physicochemical properties of lipoproteins has been scarcely studied. In a small study, polyphenols from olive oil and from thyme were associated with an improvement on lipoprotein particle atherogenic ratios and on profile distribution of lipoprotein subclasses [25]. In addition, in a sub-study of the PREDIMED trial in which lipoproteins were profiled by NMR [26], MedDiet enriched with nuts shifted lipoprotein subfractions to a less atherogenic pattern.

The aim of this study was to evaluate the effect of an intensive weight loss program, based on an energy-reduced traditional MedDiet (er-MedDiet), physical activity (PA) promotion, and behavioral support (er-MedDiet+PA), on the physicochemical properties of lipoproteins assessed by ADLT, in comparison with an energy-unrestricted MedDiet (control group) after 6 months and 1-year follow-up.

## 2. Materials and Methods

### 2.1. Study design

This is a prospective cohort study of participants from the PREDIMED-Plus trial. The PREDIMED-Plus study is an ongoing multicenter, parallel-group, randomized, single-blind clinical trial involving 6,874 participants that were recruited in 23 Spanish centers. The aim of PREDIMED-Plus study is to assess the long-term effects of an intensive weight loss program on cardiovascular events and mortality, in comparison with a MedDiet (control group) (protocol available at https://www.predimedplus.com/wp-content/uploads/2018/11/Protocolo-PREDIMED-Plus_Eng.pdf). Participants were randomly assigned, in a 1:1 ratio, to one of two groups: an intensive weight-loss intervention group or a control group. In summary, the intensive weight loss program consists of an er-MedDiet together with the promotion of physical activity and behavioral support for specific weight loss goals. The er-MedDiet intervention targeted a reduction of approximately 30% in estimated energy requirements, which represented a reduction goal of approximately 600 kcal/day [27,28]. In addition, the er-MedDiet aimed to promote better overall diet quality through the limitation of certain foods such as sugar-sweetened beverages, butter and cream, red and processed meats, added sugars, sweets, pastries and refined grains, including white bread, in favor of whole grains. Physical activity promotion included a face-to-face educational program [29] aimed at gradually increasing participants’ aerobic physical activity levels to meet at least World Health Organization (WHO) guidelines based on age, the health status of the participants [30] and static exercises to improve endurance, strength, flexibility and balance. Participants in the control group were encouraged to follow an unrestricted energy MedDiet, had biannual educational sessions on the traditional Medical Diet with ad libitum caloric intake, and received usual attention to general lifestyle recommendations [31].

The trial was registered in 2014 at the International Standard Randomized Controlled Trial registry as number 89898870 (http://www.isrctn.com/ISRCTN89898870). The primary outcome of this sub-study of the Predimed-Plus trial was to analyze the effect of an intensive weight loss program on the lipoprotein profile evaluated by ADLT. All participants provided a written informed consent form, and the study protocol and procedures were approved according to the ethical standards of the Declaration of Helsinki by all the participating institutions and the Research Ethics Committees from all recruiting centers.

### 2.2. Study subjects

The participants were community-dwelling men aged 55–75 years and women aged 60–75 years, without documented history of CVD at baseline, with a body mass index (BMI) ≥ 27 and < 40 kg/m^2^, and with at least 3 of the 5 criteria for MetS [32]. Participants were recruited and randomly allocated, in a 1:1 ratio, to either the intervention or the control group. Only those visited at the Hospital Universitari de Bellvitge (L’Hospitalet de Llobregat) were included in this study. Intervention group participants were prescribed an er-MedDiet+PA, and received personal behavioral support following the PREDIMED-Plus intervention protocol. Control group subjects were prescribed the original unrestricted-energy MedDiet and received conventional health care recommendations. The flow chart is shown in figure S1 in supplementary material.

Anthropometric measures were recorded and blood samples were collected. Overweight was defined as a BMI between 25 and 30 kg/m^2^ and obesity as a BMI ≥ 30 kg/m^2^. Dietary data were collected using a validated semiquantitative food frequency questionnaire, including 143 items commonly consumed in Spain [33]. Physical activity was measured in MET*min/week using the Regicor Short Physical Activity Questionnaire [34].

### 2.3. Methods

#### 2.3.1 Conventional lipid profile

Blood samples were taken after 12 hours of fasting during the baseline visit and then every six months for a year. The samples were collected in tubes that contained a separating gel but did not contain a coagulant (Vacuette ref: 456069). The tubes were centrifuged at 1500 g for 10 minutes (6K15 SIGMA centrifuge). The serum was immediately separated and stored at −80°C until they were analyzed.

Total cholesterol was measured by molecular absorption spectrometry at 505 and 700 nm. By the action of cholesterol esterase, cholesterol esters are separated into free cholesterol and fatty acids. The enzyme cholesterol oxidase catalyzes the reaction that transforms free cholesterol to cholest-4-en-3-one and hydrogen peroxide. In the presence of peroxidase, phenol and 4-aminophenazone, hydrogen peroxide forms a red quionimine dye. The chromatic intensity of the dye is directly proportional to the concentration of cholesterol in the sample [35, 36].

The HDL-C concentration was measured by molecular absorption spectrometry. When exposed to a detergent, non-HDL lipoproteins, including chylomicrons, VLDL and LDL form a water-soluble complex wherein the enzymatic reactions of cholesterol esterase and cholesterol oxidase are inhibited, so that only HDL particles can react with the two enzymes.

Triglycerides were measured by molecular absorption spectrometry at 505 and 700 nm. This method uses lipoprotein-lipase, glycerokinase, glycerol phosphate oxidase and peroxidase. The lipoprotein-lipase hydrolyzes the triglycerides to free fatty acids and glycerol; the latter is oxidized to dihydroxyacetonephosphate and hydrogen peroxide; subsequently the peroxide reacts with 4-aminophenazone and 4-chlorophenol to give a red dye. The intensity of the dye is directly proportional to the concentration of triglycerides present in the sample.

All the analysis were performed at Cobas c501 (Roche® Diagnostics). The reagents used in the homogenous automatized assays were Cholesterol Gen 2 (Ref: 03039773190) for Cholesterol, HDL-Cholesterol plus 3rd generation for HDL-C (Ref: 05168805190) and TRIGL Triglycerides (Ref: 08058687190) for Tg.

All analytical series were validated measuring internal controls with known concentration provided by Bio Rad Laboratories. Currently, the laboratory participate in external quality control program, Referenzinstitut für Bioanalytik, to verify the results accuracy.

The percentage of participants that were at National Cholesterol Education Program (NCEP) Adult Treatment Panel-III (ATP-III) lipid goals was evaluated [37]. These goals were: LDL-C ≤ 3.37 mmol/L, HDL-C ≥ 1.04 mmol/L, Tg ≤ 1.70 mmol/L, and Non-HDL-C ≤ 4.14 mmol/L [37].

LDL-C was estimated by the Friedewald equation and non-HDL-C was calculated by subtracting HDL-C from total cholesterol.

#### 2.3.2 Advanced Lipoprotein precipitation assays

sdLDL-C concentrations were determined using a lipoprotein precipitation method that had been adapted to clinical routine laboratory settings [38,39]. The precipitation assay was carried out 2 weeks after blood extraction. To isolate sdLDL particles, 300 uL of sample was combined with 300 uL precipitation reagent (150 U/mL heparin-Na+, catalog #H3393; Sigma-Aldrich; 90 mM MgCl_2_). The mixtures were then incubated at 37°C for 10 min, placed at 0°C for 15 min, and then centrifuged at 21913× g (14000 rpm) for 15 min at 4°C (centrifuge catalog #6K15; Sigma-Aldrich). Lipoproteins which density was <1.044 g/mL remained at the bottom of the tube, forming a yellow precipitate. The supernatant contained both HDL and sd-LDL particles which density fell between 1.044 and 1.063 g/mL. The concentration of supernatant derived HDL-C and total cholesterol were determined using a Cobas 8000 modular analyzer (Roche® Diagnostics) [40]. Since the supernatant only contained cholesterol from HDL and sd-LDL lipoproteins, the sdLDL-C concentration was estimated by subtracting the HDL-C from the concentration of total cholesterol. The reference value for the concentration of sdLDL-C is 0.04–0.47 mmol/L [38].

#### 2.3.3 Advanced lipoprotein profile by NMR spectroscopy

As previously reported, 200 μL of serum were diluted with 50 μL deuterated water and 300 μL of 50 mM phosphate buffer solution (PBS) at pH 7.4. 1H-NMR spectra were recorded at 310 K on a Bruker Avance III 600 spectrometer (Bruker BioSciences, Madrid, Spain) operating at a proton frequency of 600.20 MHz (14.1 T) [41].

Complete lipoprotein profiles were determined using the Liposcale® test. Liposcale tests are based on a 2D diffusion-ordered 1H Nuclear Magnetic Resonance (NMR) methodology to characterize lipoprotein subclasses, such as size, lipid composition, and number of particles [41]. These profiles included: 1) triglyceride and cholesterol concentrations; 2) the size and number of VLDL, LDL and HDL particles; and 3) the number of large, medium and small subclasses of VLDL, LDL and HDL particles. The particle concentrations and size were derived from the NMR signals of the univocally associated methyl lipid groups that vary between the lipoprotein subclasses.

The reference values for the advanced lipid profile are as follows: VLDL-P (24.8-50.0) nmol/L; lVLDL-P (0.70-1.18) nmol/L; mVLDL-P (2.50-5.37) nmol/L; sVLDL-P (21.7-44.1) nmol/L; LDL-P (1128-1498) nmol/L; lLDL-P (163-214) nmol/L; mLDL-P (320-513) nmol/L; sLDL-P (598-786) nmol/L; HDL-P (25.2-33.1) μmol/L; lHDL-P (0.23-0.31) μmol/L; mHDL-P (7.98-11.0) μmol/L; sHDL-P (16.7-22.4) μmol/L; VLDL-C (4.66-13.8) mg/dL; LDL-C (111-149) mg/dL; HDL-C (44.4-66.7) mg/dL; VLDL-Tg (36.4-71.4) mg/dL; LDL-Tg (12.6-19.5) mg/dL; HDL-Tg (10.3-15.4) mg/dL [42].

#### 2.3.4 Statistical analyses

A descriptive analysis of the clinical and demographic variables and lipid profiles was carried out. Categorical variables were presented as the number of cases and percentages. Continuous variables were presented as mean and 95 % confidence interval (95% CI). Normality of variables was assessed with graphs (QQ-Plot, density and standard deviations) and only the distribution of triglycerides was skewed and was log-transformed.

Differences in lipid and anthropometric variables between study groups were assessed over time using linear mixed model. To correct for potential confounding, the study group’s comparison was adjusted by sex, age, lipid-lowering treatment, and smoking status. Moreover, an interaction between study group and time was considered in order to model a differential evolution of the group effect over time. A post hoc analysis by sex was also performed by using linear mixed models adjusted by the same covariates except sex. When appropriate, estimators are shown with 95% confidence intervals. All analyses were performed with a two-sided significance level of 0.05 and conducted with the use of R software version 3.6.1 to estimate the linear mixed model the package lmer4 was used [43].

## 3. Results

The baseline clinical characteristics of study subjects are depicted in Table 1. All subjects included in the study were obese or overweight. Of these,58.4% were on lipid-lowering treatment. Conventional lipid profiles were performed during baseline visits.

**Table 1.** Descriptive analysis of demographic, clinical, and anthropometric data.

|  | Control group<br>n=95 | Intervention group<br>n=107 | *P* |
| --- | --- | --- | --- |
| Age (years) | 64.2 (63.3, 65.2) | 64.8 (63.9, 65.6) | 0.427 |
| Men (%) | 49 (51.6%) | 54 (50.5%) | 0.875 |
| Smoker (%) |  |  | 0.774 |
| Current smoker | 12 (12.6%) | 15 (14.0%) |  |
| Former smoker | 42 (44.2%) | 42 (39.3%) |  |
| Never smoker | 41 (43.2%) | 50 (46.7%) |  |
| Education (%) |  |  | 0.619 |
| Elementary (5 – 8 years) | 48 (50.5%) | 61 (57%) |  |
| Secondary (9 – 12 years) | 26 (27.4%) | 27 (25.2%) |  |
| PostSecondary (>12 years) | 21 (22.1%) | 19 (17.8%) |  |
| Hypertension (%) | 79 (83.2%) | 92 (86%) | 0.578 |
| Diabetes (%) | 29 (30.5%) | 37 (34.6%) | 0.540 |
| Dyslipidemia (%) | 76 (80%) | 82 (76.6%) | 0.563 |
| Lipid-lowering treatment (%) | 55 (57.9%) | 63 (58.9%) | 0.887 |
| Weight (kg) | 85.4 (82.8, 87.9) | 85.3 (82.9, 87.8) | 0.997 |
| Body mass index (kg/m<sup>2</sup>) | 31.9 (31.4, 32.5) | 32.0 (31.5, 32.6) | 0.754 |
| Waist circumference (cm) | 107 (105.1, 108.8) | 107 (105.3, 109) | 0.925 |
| Hip (cm) | 107 (105.6, 108.6) | 108 (106.2, 109.4) | 0.555 |
| Waist-to-hip ratio | 1.00 (0.98, 1.02) | 1.00 (0.98, 1.01) | 0.629 |
| LDL-cholesterol (mmol/L) | 3.04 (2.84, 3.23) | 3.00 (2.83, 3.16) | 0.765 |
| LDL-cholesterol ≤ 3.37 mmol/L (%) | 57 (66.3%) | 71 (68.3%) | 0.771 |
| HDL-cholesterol (mmol/L) | 1.24 (1.16, 1.31) | 1.25 (1.18, 1.31) | 0.802 |
| HDL-cholesterol ≥ 1.04 mmol/L (%) | 62 (65.3%) | 75 (70.1%) | 0.463 |
| Triglycerides (mmol/L)* | 1.73 (1.56, 1.92) | 1.57 (1.43, 1.71) | 0.152 |
| Triglycerides ≤ 1.70 mmol/L (%) | 48 (50.5%) | 60 (56.1%) | 0.430 |
| Non-HDL-cholesterol (mmol/L) | 3.81 (3.62, 4.0) | 3.49 (3.25, 3.73) | **0.045** |
| Non-HDL-cholesterol ≤ 4.14 mmol/L (%) | 63 (66.3%) | 70 (68.6%) | 0.729 |

LDL (Low Density Lipoprotein); HDL (High Density Lipoprotein); Non-HDL (Non-High-Density Lipoprotein). Data are shown as a mean and 95% confidence interval or as absolute frequency and percentage and were analyzed by the chi-square test and analysis of variance, respectively.
\*Triglycerides have been analyzed by logarithmic transformation and are expressed as antilogarithm.

During the baseline visit, the majority of MetS participants were at conventional lipid profile goals (Table 1). The percentage of participants with an LDL-C ≤ 3.37 mmol/L, with an HDL-C ≥ 1.04 mmol/L, together with a Non-HDL-C ≤ 14 mmol/L was higher in the intervention group. However, mean LDL-C and mean HDL-C were similar and mean triglyceride concentrations were slightly higher in the intervention group. Mean LDL-C in the intervention group was lower than the upper cut-off value defined by the NCEP [37].

Baseline energy, nutrients intake and Physical activity, and changes at 6 and 12 months of follow-up by intervention group are shown in table 2. As shown in Table 2, the individuals in the intervention group followed an energy-restricted diet, and a lower intake of carbohydrate and a higher intake of monounsaturated fat than the control group. Physical activity was significantly higher at 6 months in the intervention group and almost significantly higher at 12 months.

**Table 2:** Baseline energy, nutrient intake and Physical activity, and changes at 6 and 12 months of follow-up by treatment allocation.

| *Energy and Nutrients* | *Visit* | *Control group*<br>*n=95* | *Intervention group*<br>*n=107* | *P value* |
| --- | --- | --- | --- | --- |
| Energy – Kcal/day | Baseline | 2352 (2213, 2490) | 2343 (2221, 2464) | 0.923 |
|  | 6m.change | -215 (-338, -91) | -252 (-356, -148) | 0.650 |
|  | 12m.change | -250 (-378, -122) | -252 (-356, -148) | 0.982 |
| Protein - % Energy | Baseline | 17.9 (17.1, 18.6) | 17.6 (17.0, 18.3) | 0.622 |
|  | 6m.change | +0.7 (+0.03, +1.3) | +2.9 (+2.2, +3.5) | **<0.001** |
|  | 12m.change | +1.0 (+0.4, +1.7) | +2.7 (+2.0, +3.4) | **0.001** |
| Carbohydrate - % Energy | Baseline | 37.7 (36.2, 39.2) | 39.1 (37.8, 40.4) | 0.165 |
|  | 6m.change | -4.2 (-5.7, -2.7) | -6.6 (-7.9, -5.2) | **0.021** |
|  | 12m.change | -3.8 (-5.2, -2.3) | -7.0 (-8.4, -5.7) | **0.001** |
| Total fat - % Energy | Baseline | 42.1 (40.8, 43.5) | 40.4 (39.2, 41.6) | 0.058 |
|  | 6m.change | +3.6 (+2.1, +5.1) | +4.2 (+2.9, +5.5) | 0.522 |
|  | 12m.change | +2.5 (+1.1, +3.9) | +4.7 (+3.3, +6.2) | **0.028** |
| SFA - % Energy | Baseline | 10.9 (10.4, 11.3) | 10.2 (9.8, 10.6) | **0.035** |
|  | 6m.change | -0.7 (-1.2, -0.2) | -1.2 (-1.6, -0.8) | 0.120 |
|  | 12m.change | -1.3 (-1.8, -0.8) | -0.7 (-1.1, -0.3) | 0.070 |
| MUFA - % Energy | Baseline | 21.8 (20.8, 22.8) | 20.6 (19.8, 21.4) | 0.057 |
|  | 6m.change | +4.0 (+2.8, +5.1) | +5.8 (4.9, +6.8) | **0.012** |
|  | 12m.change | +3.3 (+2.2, +4.4) | +5.9 (+4.8, +6.9) | **0.001** |
| PUFA - % Energy | Baseline | 7.2 (6.8, 7.6) | 7.1 (6.7, 7.4) | 0.638 |
|  | 6m.change | +1.1 (+0.7, +1.5) | +1.4 (+1.0, +1.9) | 0.204 |
|  | 12m.change | +1.2 (0.7, +1.6) | +1.6 (+1.2, +2.0) | 0.170 |
| Fiber – g/day | Baseline | 25.2 (23.7, 26.6) | 24.5 (23.0, 26.1) | 0.551 |
|  | 6m.change | +2.7 (1.0, +4.5) | +5.7 (+3.8, +7.5) | **0.026** |
|  | 12m.change | +2.8 (+1.0, +4.3) | +4.9 (+3.2, +6.6) | 0.067 |
| Cholesterol – mg/day | Baseline | 399 (375, 424) | 396 (373, 419) | 0.852 |
|  | 6m.change | -25.6 (-49.0, -2.2) | -29.1 (-51.2, -6.9) | 0.833 |
|  | 12m.change | -27.8 (-52.5, -3.1) | -13.2 (-38.0, +11.6) | 0.413 |
| Physical activity –<br>MET*min/week | Baseline | 2336 (1957, 2715) | 2614 (2240, 2987) | 0.303 |
|  | 6m.change | +404 (45, 763) | +1263 (847, 1681) | **0.002** |
|  | 12m.change | +569 (8, 1129) | +1242 (743, 1741) | 0.076 |

Data are means (95% Confidence Intervals) and analysed by 1-way analysis of variance. 6m.change (6 months change versus baseline); 12m.change (12 months change versus baseline); SFA (saturated fatty acids); MUFA (monounsaturated fatty acids); PUFA (polyunsaturated fatty acids); MET (metabolic equivalent).

Advanced lipoprotein profiles were also determined during baseline visits (see supplementary material Table S1). The proportion of LDL-C content in small LDL particles and the proportion of particles content in each lipoprotein subclass were calculated as percentages. The mean sdLDL-C was higher than the upper cut-off value of the reference interval [38]. The predominant subclasses of LDL particles were sdLDL-P, which accounted for 61.2% of the total LDL particle number (LDL-P), and sHDL-P were the dominant subclass of total HDL particle number (HDL-P), representing 66.7% (supplementary material Table S1). These results are consistent with the phenotype of MetS dyslipidemia.

### Effect of MedDiet and er-MedDiet+PA based intervention on anthropometric characteristics and lipid profile

Predictor’s coefficients and *p*-values were used to quantify the predictive effect of the MedDiet and er-MedDiet+PA on changes in anthropometric characteristics and lipid profiles that occurred between the baseline and follow-up visits. In Table 3 the more relevant variables from mixed model analysis are shown. Results of the intervention at 6 and 12 months are adjusted according to group, time, interaction group and time, sex, age, the administration of lipid-lowering treatments, and smoking status.

**Table 3.**
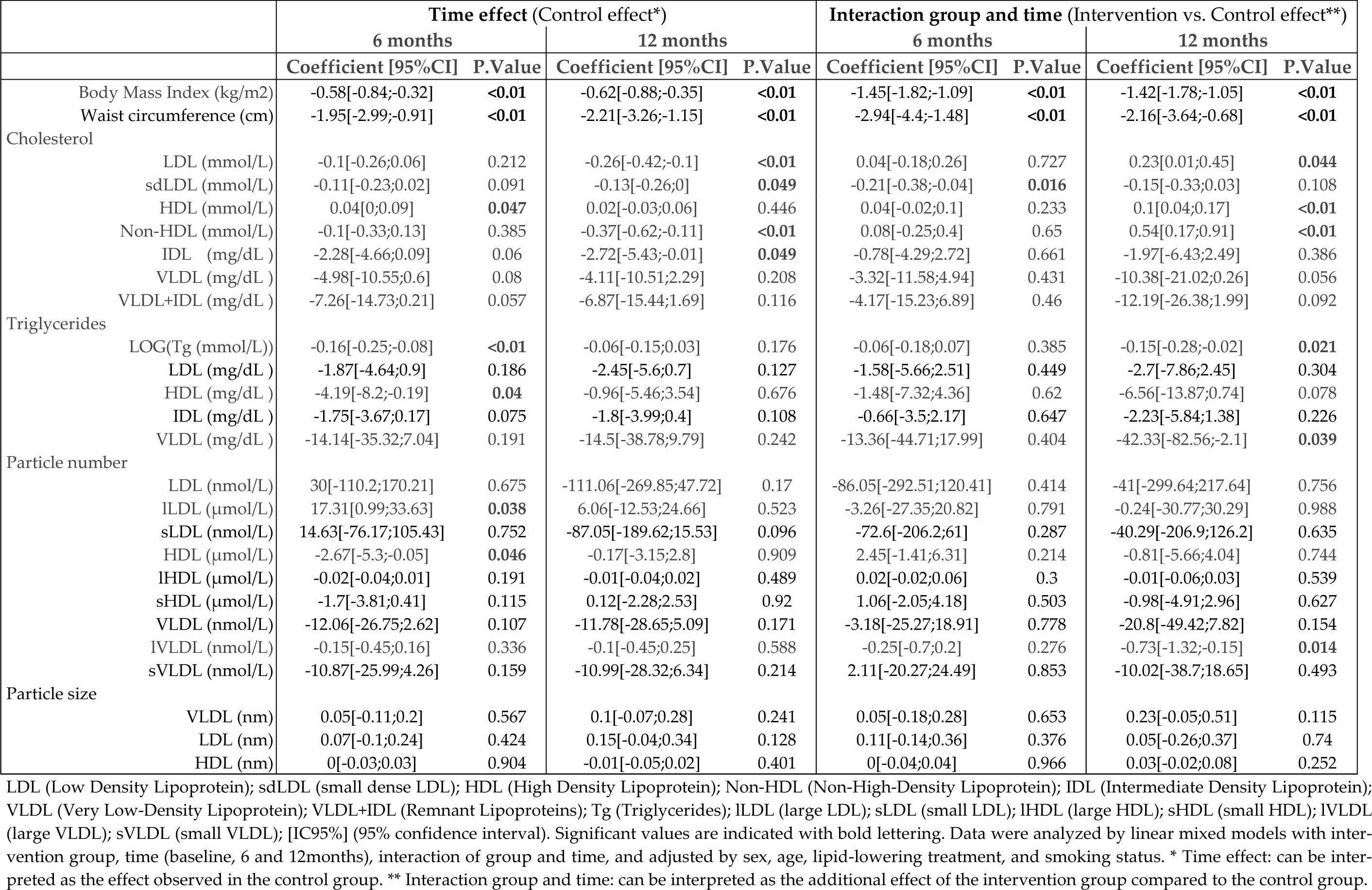
Effects of MedDiet and er-MedDiet+PA based intervention on lipid and anthropometric variables observed during follow-up visits.

#### Anthropometric variables

Participants in the MedDiet control group sustained an improvement in all anthropometric variables at 6 months and 1 year. Table 3 shows that at 6 months of follow-up, the variables BMI and waist circumference decreased significantly in the final adjusted model. Weight decreased 1.7 kg at 6 months and 1.7 kg at 12 months. A further weight loss of 3.9 kg (*p*<0.01) and 3.9 kg (*p*<0.01) and a reduction in waist circumference of 2.9 cm (*p*<0.01) and 2.2 cm (*p*<0.01) at 6 months and 12 months respectively, was predicted in the er-MedDiet+PA intervention group in comparison with the control group (*p*<0.01) (Figure 1).

**Figure 1.**
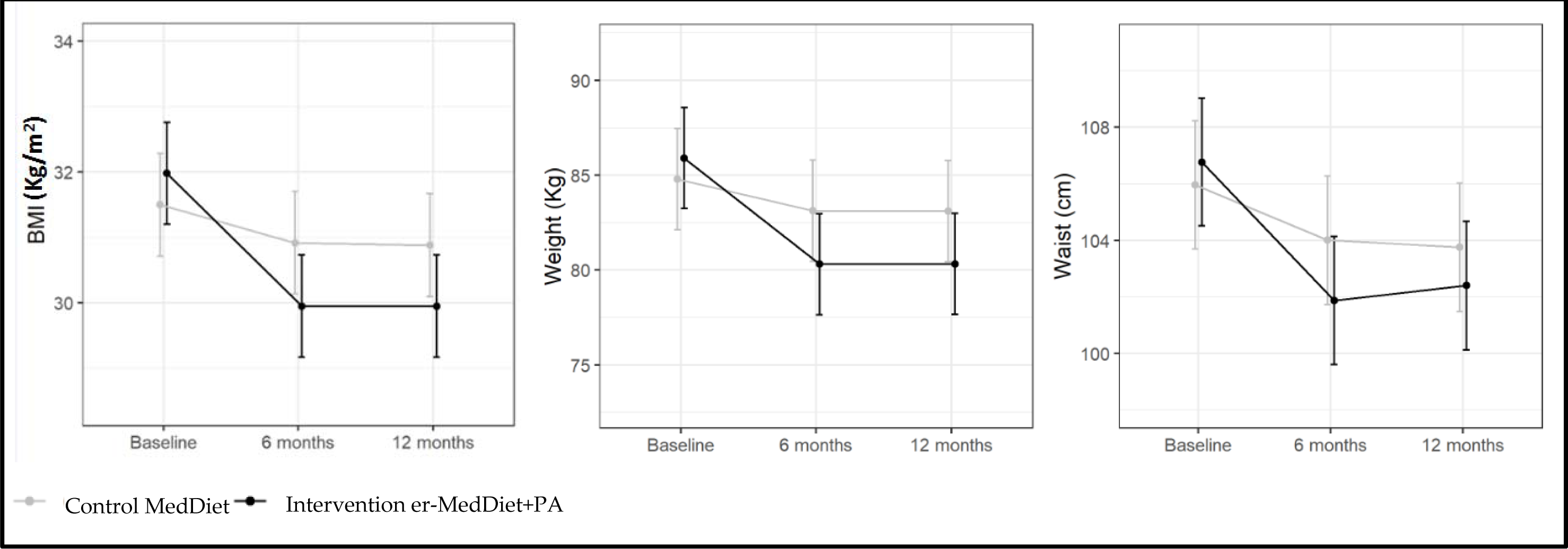
Effect of MedDiet and er-MedDiet+PA on anthropometric variables at 6 and 12 months.

#### Lipid profile

The effect of traditional MedDiet on the lipid panel evaluated by means of NMR is shown in the first two columns of table 3. The control group shows an increase in HDL-C and a decrease in serum Tg concentrations. These results were significant at 6 months (p= 0.05, and p<0.01, respectively). LDL-C and non-HDL-C concentrations decreased at 12 months 0.26 mmol/L (p<0.01) and 0.37 mmol/L (p<0.01), respectively. The additional effect of er-MedDiet+PA on the ADLT measurements is shown in the last two columns of table 3. In comparison with the control group, the er-MedDiet+PA-based intervention program led to a significant further reduction of Tg (p=0.021) (Supplementary material Figure S2) and to an increase of 0,1 mmol/L of HDL-C (p<0.01) (Supplementary material Figure S5). Also and increment in LDL-C and Non-HDL-C concentration was detected in the intervention group (p= 0.05, and p<0.01, respectively).

#### Advanced lipoprotein tests

In the control group sdLDL-C and IDL-C decreased 0,13 mmol/L (*p*=0.05) and 2.72 mg/dL (*p*=0.05) at 12 months and HDL-Tg and HDL-P decreased at 6 months (4.19 mg/dL, *p*=0.04 and 2.67μmol/L, *p*=0.046, respectively). In comparison with the control group, in the intervention group a further decrease in sdLDL-C and sdLDL-C/LDL-C (%) of 0.21 mmol/L and 9.23% was observed at 6 months (p=0.016, p<0,01, respectively, and also a decrease in VLDL-Tg and large VLDL particles (lVLDL-P) of 42.33 mg/dL (*p*=0.039) and 0, 73 nmol/L (*p*=0.014), respectively at 12 months. No changes in LDL-P were observed. These results are shown in table 3 and Figure 2. Effect on VLDL-C, VLDL-P, LDL and HDL advanced lipid profile results are shown in supplementary material Figures S3-S5. Analyzes of all the data included in Table 3 were also performed separately by sex and non-clinically relevant differences were observed (supplementary material, tables S2 and S3).

**Figure 2.**
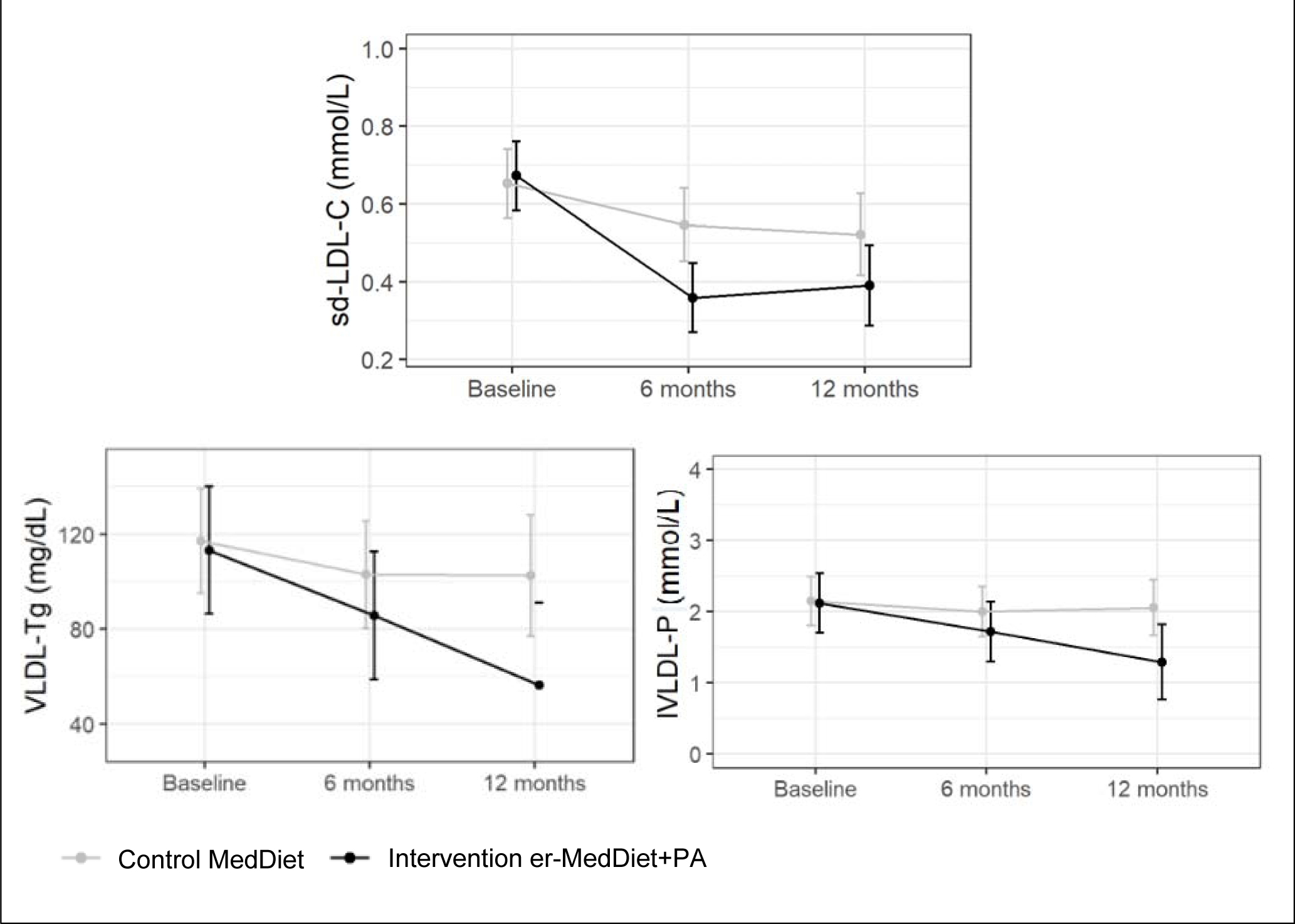
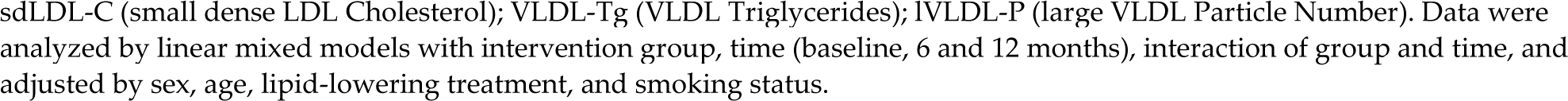
Effect of MedDiet and er-MedDiet+PA on sdLDL-C, VLDL-Tg and lVLDL-P at 6 and 12 months.

## 4. Discussion

The results of the original PREDIMED trial demonstrated that a non-energy restricted MedDiet reduced the incidence of CVD among participants at high cardiovascular risk who were mostly overweight/obese [14]. In PREDIMED the energy-unrestricted MedDiet intervention improved the lipid profile and led to small reductions in waist circumference and body weight [44,45]. On the other hand, in the last decades lipoprotein assays have been developed which are able to detail the composition of lipoproteins and can distinguish between their subclasses [41]. These advanced methods can be used to update our understanding of the effects of diet and lifestyle on lipid metabolism.

We compared the effects of an energy-unrestricted traditional MedDiet and an er-MedDiet+PA intervention on body weight and lipid profiles, including ADLT, in MetS patients enrolled in the PREDIMED-plus trial at Hospital Universitari de Bellvitge. All patients were overweight or obese and 73.8% were treated with lipid-lowering drugs (Table 1). At baseline, most participants had not overt dyslipidemia judging from their conventional lipid profiles (Table 1) and, although their mean LDL-C was lower than the upper limit defined by the NCEP [37], the mean sdLDL-C was higher than the upper limit of the reference interval [33]. In addition, their mean LDL-P number was higher than the treatment target value, as defined by the American Association of Clinical Endocrinologist’s [46]. These data illustrate that changes in the concentration of sdLDL-C and LDL-P not always go in parallel with changes in LDL-C. Furthermore, sdLDL-C and LDL-P remained abnormally elevated even in patients classified as low risk by their LDL-C level. Importantly, the dominant subclasses of LDL and HDL particles were found to be sLDL-P (61.2% of all LDL-P) and sHDL-P (67,7% of all HDL-P), respectively (Table 3), values that are consistent with the MetS phenotype. MetS subjects are thought to have a smaller mean HDL particle size, and it has been hypothesized that this alteration is linked to inflammation [47]; however, this concept needs to be supported by further research [48].

### Anthropometric variables

Adoption of the (energy-unrestricted) MedDiet led to an improvement in all anthropometric variables at 6 months and at 1 year of intervention (Table 3). These results are consistent with those observed in the PREDIMED trial. The model predicted a weight loss of 1.7 kg and a reduction in waist circumference of 1.9 cm at 6 months (*p*<0.01). Importantly, these results remained significant at 12 months. In comparison, the er-MedDiet+PA intervention had a remarkable impact on weight, waist and hip measurements (*p* < 0.01). In fact, the adjusted model predicted an additional reduction of 3.9 kg, 2.9 cm and 1.8 cm, respectively (Table 3) at 6 months, and the effect persisted at 12 months. Expectedly, the er-MedDiet+PA intervention program was more effective than the MedDiet without energy restriction to achieve the weight loss targets. It has been shown repeatedly that overweight or obese persons following an er-MedDiet with or without enhanced PA lose weight and this holds as long as the diet has energy curtailed [49].

### Conventional lipid profile

After 6 months of MedDiet, a decrease in triglycerides and an increase in HDL-C concentrations was observed in both groups. However, after 12 months, these differences decreased in magnitude and lost statistical significance in the control group, while they remained significant in the intervention group. These findings are consistent with those reported in a recent meta-analysis of RCTs of MedDiet for MetS components (9). Regarding the er-MedDiet+PA intervention, raised HDL-C and reduced Tg concentrations were observed at 12 months compared with the conventional MedDiet, highlighting the importance of the intensive intervention on these 2 components of the lipid triad. These results demonstrate that the MedDiet combined with traditional health care has a beneficial effect on LDL-C and non-HDL-C levels beyond what is accomplished with lipid lowering treatment alone. Furthermore, the er-MedDiet+PA led to greater improvement of lipid variables linked to MetS, such as Tg and HDL-C. Despite this improvement in Tg and HDl metabolism, and despite the weight loss, an increment on LDL-C and Non-HDL-C concentration was seen.

### Advanced lipid profile

With respect to ADLT measurements, sdLDL-C, IDL-C and HDL-Tg, but not LDL-P, significantly decreased in response to the traditional MedDiet. sLDL-P number did not change significantly, but a non-significant negative regression coefficient was observed for this variable at 12 months. In addition, the cholesterol content in sdLDL decreased significantly at 12 months. It has been well established that caloric restriction and exercise have a favorable effect on triglyceride and lipoprotein metabolism [50]. However, the effect of the traditional MedDiet on LDL-P concentration and subclass distribution has been scarcely studied. In previous studies with MedDiet including subjects with metabolic syndrome, increases in LDL size and a favorable redistribution of cholesterol among the different LDL particles were observed [26,51,52], whereas in a study of healthy subjects the MedDiet had no effect [53]. Although in the current study, LDL-P did not change, the MedDiet was associated with an improvement in LDL particle composition and sdLDL-C was reduced. In addition, HDL-Tg was decreased at 6 months; as far as we know, this is the first study that has demonstrated this effect. Previous studies have reported higher HDL-Tg in patients with carotid atherosclerosis in association with other biomarkers linked to the MetS [54]. A decrease in HDL-P was also observed. A reasonable explanation for these results is that a change in HDL composition had occurred. However, no significant changes in HDL size or HDL subclasses were observed. HDL-P and sHDL-P are inversely related to CVD mortality and even all-cause mortality in patients with coronary artery diseases [55]. A nearly significant decrease in IDL-C and IDL-Tg (major determinants of pro-atherogenic remnant lipoproteins) was also observed after the conventional MedDiet.

In comparison with the standard MedDiet, the er-MedDiet+PA-based intervention led to a significant reduction of sdLDL-C and the sd-LDL-C/LDL-C ratio, but no changes of LDL-P. In accordance with our results, in a recent randomized controlled feeding trial no decrease in LDL-P concentration was found in individuals following a low-carbohydrate diet who had lost weight [56]. Expectedly, the er-MedDiet+PA intervention had a beneficial effect on variables related to Tg metabolism at 12 months. VLDL-C was reduced with borderline statistical significance (*p*=0.056) and a similar trend was observed for the VLDL-C+IDL-C concentration (*p=*0.092). The er-MedDiet+PA intervention also induced a decrease in VLDL-Tg and, as a consequence, lVLDL-P were also reduced and LDL particle size increased, changes that are recognized as anti-atherogenic [57]. HDL-triglyceride also tended to decrease with the er-MedDiet+PA intervention. It should be noted that a moderate decrease in carbohydrate consumption and a moderate increase in monounsaturated fat consumption was observed in the intervention group compared to the control group, and these changes could have influenced the lipid responses. Low-or very-low CHO diets (so-called ketogenic diets), which usually are reciprocally enriched in fat, are superior to low-fat diets in improving cardiometabolic risk due to a TG lowering and HDL-C raising effect, with only a negligible effect on LDL-cholesterol [58]. Also, there is ample clinical trial evidence that such CHO-restricted dietary interventions increase LDL peak particle size and decrease the numbers of total and small LDL particles [59]. Thus, diets lower in CHO and higher in fat such as the er-MedDiet used in PREDIMED-Plus have the potential to improve both the standard and the advanced lipid profile. High-fat, carbohydrate-restricted diets improve atherogenic dyslipidemia and insulin resistance [37,45–48] because carbohydrate consumption increases hepatic triglyceride synthesis and induces insulin secretion that leads to inhibition of lipolysis, enhanced delivery of fatty acids for hepatic esterification, and overproduction and secretion of large triglyceride-rich VLDL particles. In addition, other potential mechanisms may be involved in the interaction between MedDiet and the effects of statins on lipid metabolism, i.e., the reduction in plasma insulin observed with a carbohydrate restricted MedDiet may decrease the expression of the HMG-CoA reductase gene, with ensuing lower secretion of very low-density lipoprotein (VLDL)-cholesterol, which, in turn, would account for the reduction in LDL-C [60].

These large VLDL exchange triglyceride for cholesteryl ester in both LDL and HDL. Triglyceride-rich LDL is a preferred substrate for hepatic lipase that favors the production of sd-LDL particles [61,62], while triglyceride enrichment of HDL correlates inversely with the particles’ functionality [63] and is a good marker of cardiometabolic risk [54]. These changes in lipid metabolism are observed even in the absence of significant weight loss [64]. Aerobic exercise improves atherogenic dyslipidemia, among other mechanisms by increasing lipoprotein lipase activity and decreasing fasting and post-prandial serum triglycerides [65].

Although the er-MedDiet-PA intervention had an impact on lipid variables linked to Tg, a similar effect was not observed for variables linked to cholesterol. While LDL-C and Non-HDL-C levels decreased in the standard MedDiet group, they increased in the er-MedDiet+PA group. This suggests that the cholesterol-lowering effect was related to the MedDiet dietary regime alone, while added energy restriction and exercise in the er-MedDiet+PA intervention counteracted this effect. In some studies, it has been observed that LDL-C increases after a session of moderate-intensity exercise [66] or when adhering to a low-calorie diet [67]. Whether energy restriction affects LDL-C levels is controversial. Thus, there is evidence in favour of modest LDL-cholesterol lowering in response to weight loss [68]. However, there are notable exceptions from seminal studies in which weight loss was unassociated with LDL-cholesterol changes [69,70], including preliminary evidence from the PREDIMED-Plus study [71]. Beyond weight loss, macronutrient changes in the diets (i.e., saturated fatty acids) may explain these discrepancies.

Our study has some limitations. Participants in the PREDIMED-Plus trial are predominantly older white Spanish individuals with overweight or obesity harboring the MetS, which limits the generalizability of the results to other populations. Despite this, one strength of this study lies in the homogeneity of the cohort that increases the internal validity of the findings by avoiding confounding factors related to socioeconomic status or educational level. We have also controlled for other possible confounders using multivariate statistical models. Another strength of our study is the longitudinal analysis carried out in a homogeneous and sizable cohort. Also all participants were well characterized with many clinical and laboratory variables related to metabolic syndrome including dietary components and physical activity. A further strength is the use of ADLT, for lipid and lipoprotein analyses, allowing for a broad and comprehensive characterization of the spectrum of plasma lipoprotein species species and their response to er-MedDiet+PA. Finally, the design of the statistical study allowed us to obtain reliable results since many possible confounders were considered.

## 5. Conclusions

In this sub-study of a lifestyle intervention trial, we compared the effects of MedDiet alone and an er-MedDiet+PA intervention on the lipoprotein subclass profiles of MetS patients. The MedDiet alone improved lipoprotein composition with a reduction in HDL-Tg, and sdLDL-C, and an increase of ILDL particles, while the er-MedDiet+PA intervention led to an improvement of in triglyceride metabolism, i.e., a decrease in VLDL-Tg, VLDL-P and lVLDL-P. It’s needs to be highlighted that these changes in triglyceride metabolism are associated with an improvement in LDL composition and functionalism. Thus, although no changes on LDL-P concentration were observed, VLDL-P with a higher content of Tg decreased, and this change leads to the formation of less atherogenic LDL-P. Further studies with longer follow-up must be done to evaluate the effect of energy restricted diet on LDL-P concentration.

Both an energy-unrestricted MedDiet and er-MedDiet+PA intervention are promising strategies for reducing sdLDL-C by optimizing different lipid metabolic pathways. These outcomes are important because sdLDL-C is more atherogenic than lLDL-C. The combination of the MedDiet with a negative energy balance achieved by diet and physical activity has proven to be an effective approach for improving triglyceride-rich lipoprotein metabolism and reducing the concentration of atherogenic lipoproteins. As the prescription of these diets must be done by experienced professionals and is time-consuming further studies on their cost effectiveness are warranted. Also the evaluation of the effects of the lipoprotein subclass changes induced by these diets on CVD and all-cause mortality.

Our results illustrate how lipid-lowering treatment alone without lifestyle changes in patients with metabolic syndrome is not enough to reduce cardiovascular risk in patients with metabolic syndrome.

## Data Availability

The study database is available upon request addressed to the authors of the manuscript

## Acknowledgements

The authors thank CERCA Program/Generalitat de Catalunya for providing institutional support. and Department of Biochemistry, Molecular Biology and Biomedicine, Autonomous University of Barcelona (UAB), Barcelona, Spain. CIBEROBN is an initiative of ISCIII, Spain.

## Author Contribution

Conception, B.C and B.F; Acquisition, X.P, M-F-M, V.E-L, B.C and B.F; Analysis, B.C, C.T, B.F, X.P and E.C; Interpretation, B.C; B.F, E.C, M.F-M, V.E-L, J.S-S, M.F, E.R and X.P; Resources, B.C and X.P; Writing—original draft preparation, B.C and B.F; Writing—review and editing, E.C, E.R, J.S-S, M.F, T.R-M, B.C, B.F and X.P. All authors have read and agreed to the published version of the manuscript.

